# Ethnic inequalities in loneliness in Britain during the COVID-19 pandemic: Evidence for Equality National Survey (EVENS)

**DOI:** 10.1101/2025.04.02.25325107

**Authors:** Zeng-Hui Ma, Patricia Irizar, Aradhna Kaushal

**Affiliations:** Department of Epidemiology & Public Health, University College London, London, UK; Department of Psychology, Liverpool John Moores University, Liverpool, UK

**Keywords:** Ethnic inequalities, Loneliness, Mental health, COVID-19, Epidemiology

## Abstract

Ethnic minority populations faced a disproportionate impact of the COVID-19 pandemic. Loneliness, a significant public health issue, was exacerbated during the pandemic. Most previous studies used aggregated ethnic groups and overlooked underrepresented groups. This study explored ethnic inequalities in loneliness across 21 disaggregated ethnic groups in Britain during the COVID-19 pandemic. It used cross-sectional data (February to November 2021) from the Evidence for Equality National Survey (EVENS), with 14215 participants aged 18-75 from 21 ethnic groups in Britain. Weighted logistic regression models examined ethnic inequalities in loneliness across 21 disaggregated and 6 aggregated groups, both unadjusted and adjusted for socio-demographics. This study found higher odds of loneliness among most ethnic minority groups, with nuanced differences that were overlooked in aggregated analyses. Socio-demographics explained disparities for some ethnic groups, while differences in others remained after adjustment, suggesting additional factors driving these differences.

## Introduction

Ethnic minority populations were disproportionately affected by the COVID-19 pandemic, with reported higher rates of infection, severe illness, and mortality compared to the ethnic majority population (Irizar, Pan, et al. 2023). In England, higher risks of COVID-19-related infections, hospitalization, intensive care unit (ICU) admission, and death were observed in the South Asian, Black, Mixed, and Other ethnic groups compared to the White population (Mathur et al. 2021). The existing socioeconomic and health inequalities that affect ethnic minorities were exposed and further exacerbated by the COVID-19 pandemic, sparking social debates and policy agendas during and beyond the pandemic (Salway et al. 2020).

Loneliness has long been a health risk (Cacioppo 2018), linked to a range of negative physical and mental outcomes, including increased risk of cardiovascular conditions (Hawkley et al. 2006), impaired cognitive performance (Wilson et al. 2007), and depression (Cacioppo et al. 2006). During the pandemic, loneliness was more widely recognized as a significant public health issue, as individuals experienced rapid reduction of usual in-person social contact (Cénat et al. 2022). A UK longitudinal study indicated that loneliness experienced by young adults closely followed the severity of lockdown restrictions from 2020 to 2021 (Kung, Kunz, and Shields 2023). Several conceptual models were developed to illustrate the potential pathways contributing to ethnic health disparities during the pandemic, highlighting racism as a key driving force affecting multiple layers of social determinants within these pathways (Nazroo and Becares 2020; Irizar, Kapadia, et al. 2023).

The extent of ethnic disparities in loneliness is unclear in previous literature, partly due to the use of different covariates and aggregated ethnic group categories. One study using unadjusted analyses found that almost all ethnic minority groups-African, Chinese, Caribbean, Pakistani, and Bangladeshi-except the Indian group in the UK had a higher prevalence of loneliness compared to the White British group (Victor, Scambler, and Bond 2008). Other studies also found that loneliness prevalence among ethnic minorities was higher in unadjusted analyses, but when correcting for other factors such as poorer health conditions, lower socioeconomic status, and perceived discrimination, the inequalities were reversed, in many cases putting the majority group at higher risk of loneliness (Fokkema and Naderi 2013; Visser and El Fakiri 2016).

Several studies have examined ethnic inequalities in loneliness during the COVID-19 pandemic. Longitudinal data from the UK Household Longitudinal Study revealed that compared to White British group (18%), White Irish and Indian individuals were more likely to report loneliness (28% and 24%), while Chinese or other Asian, and Black, African, Caribbean or Black British individuals were less likely to report it (10% and 11%) (ONS, 2020). Another UK longitudinal study analyzing data collected before and during the first lockdown period found the Mixed ethnic group had nearly double the rates of loneliness (15.1%-16.9%) compared to other ethnic groups (White, Asian, Black, and Other groups, 5.3%-9.6%) (Niedzwiedz et al. 2021). Nevertheless, despite the large sample sizes in many of those studies, they are limited by the use of aggregated ethnic groups, which could mask potentially heterogeneous loneliness experiences among ethnic subgroups. This limitation was also present in studies with limited sample size in ethnic minorities, which prevented further subgroup disaggregation (O’Connor et al. 2021). Additionally, underrepresented groups, such as Gypsy/Traveller/Roma, who are often known to have the worst health outcomes of all (Yin-Har Lau and Ridge 2011), were often overlooked, leading to potential biases.

The current study utilized a large cross-sectional dataset across 21 ethnic groups (aged 18-75) in Britain from the Evidence for Equality National Survey (EVENS) to examine ethnic inequalities in loneliness during the COVID-19 pandemic. Both disaggregated and aggregated ethnic analyses were conducted to examine how group aggregation might affect the detection of ethnic differences in loneliness. To understand the impact of various socio-demographic factors on ethnic disparities in loneliness, the Dahlgren-Whitehead rainbow of health determinants (Göran and Whitehead 1991) was adopted to understand what might be driving differences in loneliness between ethnic groups.

## Methods

### Study design and study sample

This study used cross-sectional data from the EVENS which is the largest survey documenting the experiences of ethnic minority people in Britain during the COVID-19 pandemic (Finney et al. 2023). EVENS was conducted between February and November 2021 using a non-probability survey design to recruit more ethnic minority groups and larger samples of them. Eligible participants were individuals aged 18 or older and living in Scotland, Wales, or England. Several strategies were employed for the recruitment purpose, including an open link survey, survey panels, snowballing, and community-based interviews. Daily monitoring of survey responses was conducted to ensure that the desired sample size was achieved for each population group, based on age, sex, and region (Finney et al. 2023).

### Survey weights

As EVENS is a non-probability sample, it was necessary to use weights to adjust for selection biases and coverage biases. To calculate statistical adjustment weights, the propensity scores were first obtained using statistical modelling on both the EVENS sample and two probability-based reference samples (Annual Population Survey 2019 and 2020, and European Social Survey rounds 8 and 9). The predicted probabilities of participants were then used to create pseudo-design weights. After this, post-stratification adjustment weights were developed using age group, sex, region, and ethnic group as weighting classes to calibrate the sample to population benchmarks (Finney et al. 2023).

### Outcome variable

Experiences of loneliness were quantified by the self-completed UCLA 3-item Loneliness Scale (Hughes et al. 2004). Response for each question ranged from “Hardly ever or never” (1) to “often” (3), which gave a total score ranging from 3 to 9 (see supplementary material for details). The cutoff was set at 6, with scores equal to or larger than 6 indicating presence of loneliness, aligning with previous analyses on loneliness using EVENS dataset (Finney et al. 2023).

### Exposure variable

The EVENS dataset comprised 21 disaggregated ethnic groups as shown in Table 1. To be comparable with previous research, those ethnic groups were aggregated into 6 groups as used in 2021 UK Census (ONS, 2021), specifically: White British; White minority (Eastern European, Gypsy/Traveller, Irish, Roma, Any other White background); Mixed or multiple ethnic groups (White and Asian, White and Black African, White and Black Caribbean, Any other mixed/multiple background); Asian (Bangladeshi, Chinese, Indian, Pakistani, Any other Asian background); Black (African, Caribbean, Any other Black background); and Other ethnic group (Arab, Jewish, and Any other ethnic group).

**Table 1.**
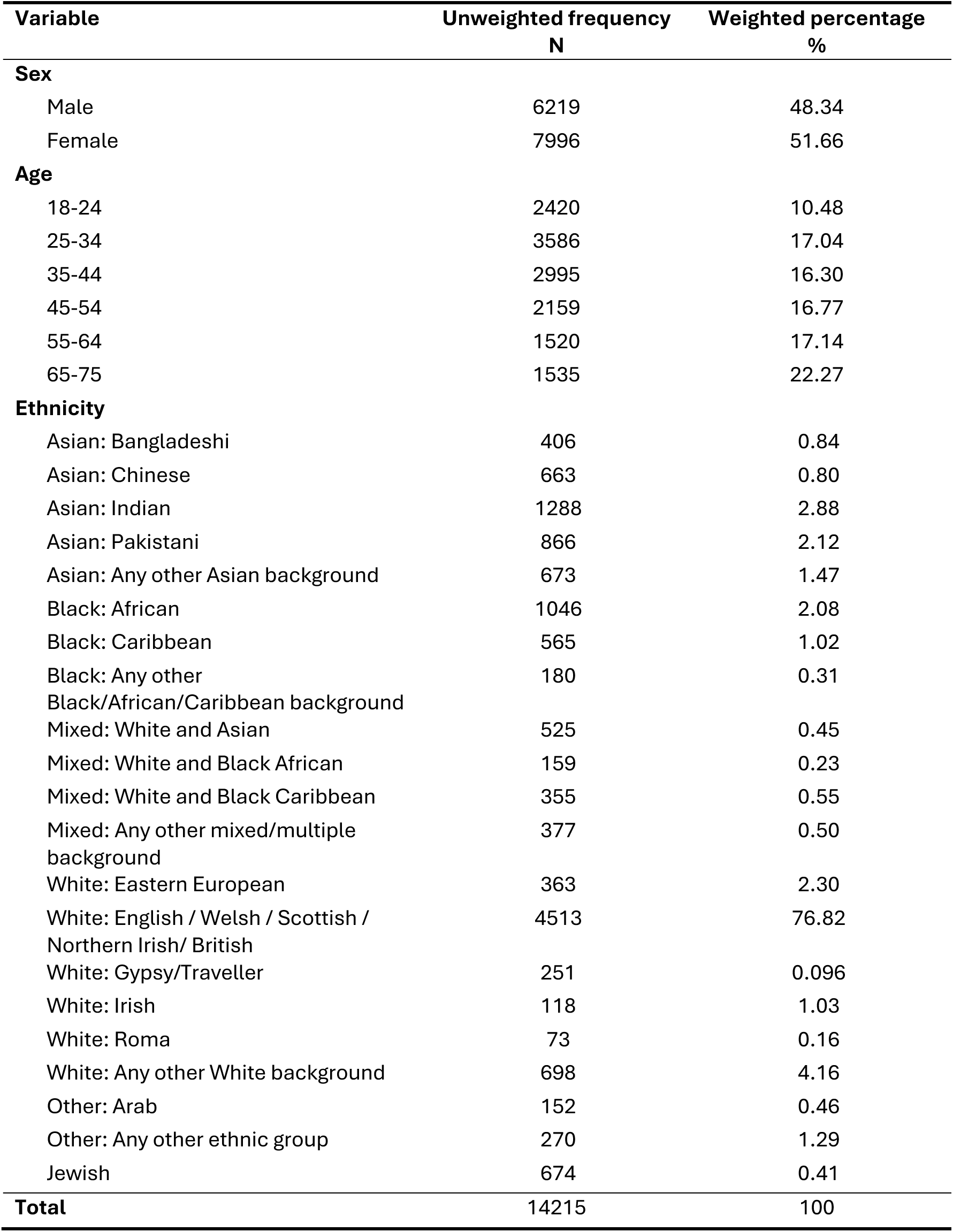
Sample characteristics (Unweighted frequency and weighted percentage, N = 14215).

### Covariates

This study included covariates previously associated with loneliness during COVID-19, as identified in earlier research (McQuaid et al. 2021; Pierce et al. 2020). The covariates considered were age, sex, previous COVID infection, marital/partnership status, household generational structure, financial concerns, region of residence, education, and employment status (see supplementary material for details).

### Missing data

The EVENS dataset comprised a total of 14221 participants. Out of the whole sample, 121 participants abandoned the online survey after completing more than two-thirds of the questions. Nearest-neighbour random hot deck imputations were conducted on 115 cases and 6 were dropped due to not completing the minimum number of questions required for imputation. This resulted in 14215 cases released on the UK Data Service. Further details were described elsewhere (Ipsos, 2023).

For the outcome variable loneliness, 3.90% of participants selected “prefer not to say” for at least one item. Person level mean imputation was applied for those with complete data on 2 out of 3 items by averaging the scores of the available 2 items, resulting in 13967 cases analyzed because other participants answered “prefer not to say” for all, or 2 out of 3 of the loneliness questions (see Figure S1 for the participant flow diagram). A sensitivity analysis was conducted using complete case analysis through listwise deletion, excluding 3.90% of cases and leaving 13660 cases in the analysis. Fewer than 5% of participants responded with “don’t know/prefer not to say” across covariates, which were included as a separate category, and no missing data were present in covariates.

### Statistical analysis

Statistical analyses were conducted using STATA 18.0. Step-by-step weighted logistic regression models were conducted based on the “Rainbow” model of the health determinants (Göran and Whitehead 1991) as shown in Figure 1. The full list of variables used can be seen in table S2. Models were constructed using the following steps: firstly only loneliness and ethnicity were included in the model, without adjusting for any covariates (model 1); then adding covariates of age, sex, and previous COVID infection to the model on individual level (model 2); adding covariates of marital/ partnership status and household generational structure on social and household level (model 3); adding covariates of financial concerns, region of residence, education, and employment status on general socio-economic and environmental level (model 4). These analyses were run on both 21 and 6 ethnic groups to compare results of aggregated and disaggregated group analyses. A sensitivity analysis was conducted using weighted linear regressions to explore ethnic differences on continuous outcomes. The variables used in the logistic regression were checked for collinearity (see supplementary material). Age was included as a categorial covariate due to the violation of linearity assumption. Significance level was set at alpha = 0.05 and 95% confidence intervals were used.

**Figure 1.**
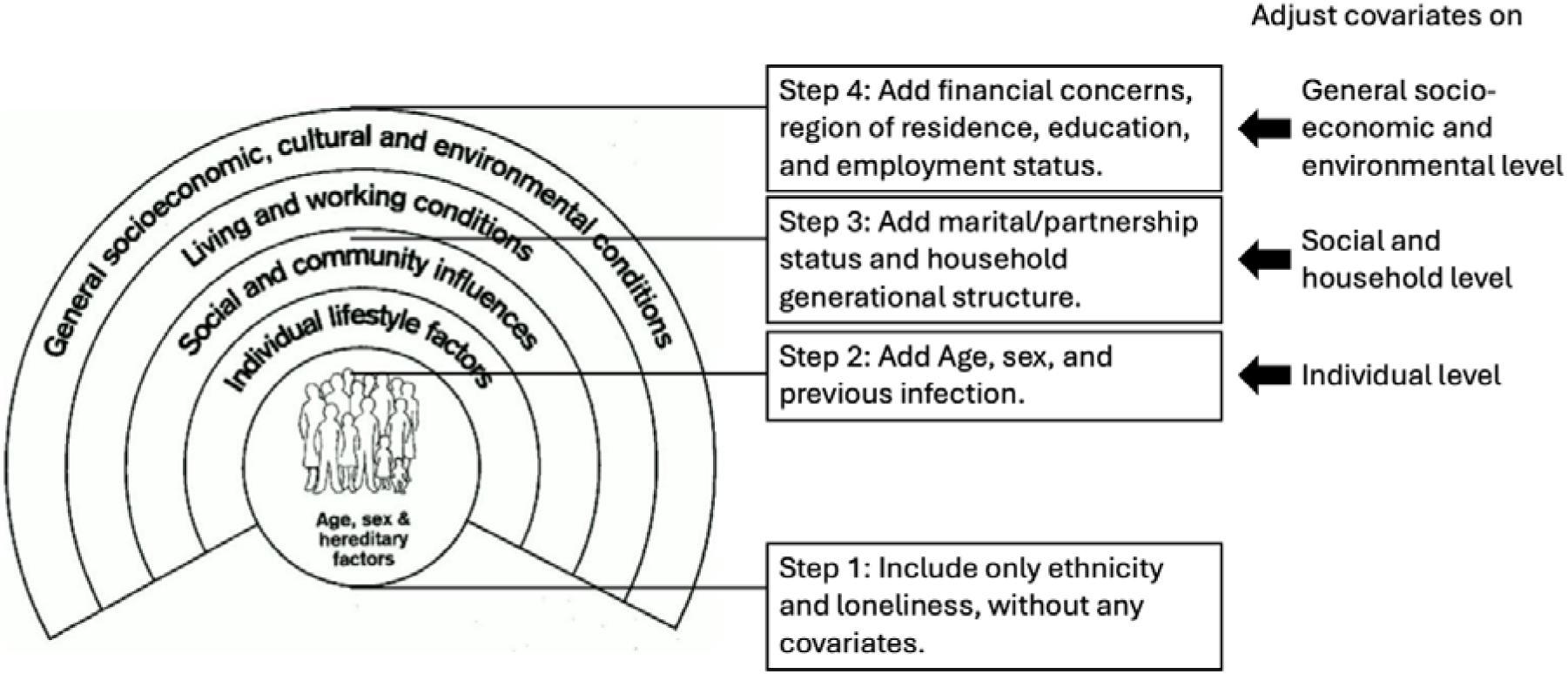
Step-by-step covariate adjustment approach based on rainbow model of health determinants, with covariates adjusted on three different levels separately.

### Ethical considerations

This study utilized the public dataset EVENS dataset accessed via the UK Data Service. The EVENS study received ethical approval from the University of Manchester Research Ethics Committee and was conducted in compliance with the Declaration of Helsinki. No additional ethical approval was necessary for this secondary analysis.

## Results

### Sample characteristics

The sample comprised 14215 participants. Unweighted frequencies and weighted percentages for ethnicity, age, and sex are shown in Table 1 (sample characteristics for other variables are shown in Table S1).

### Ethnic inequalities in loneliness across 21 ethnic groups

Compared to the White British group (Table 2), 11 out of 20 ethnic minority groups had significantly higher odds of loneliness in the unadjusted model (Model 1), including Bangladeshi (ORs: 1.46, 95% CI: 1.03 to 2.06), Indian (ORs: 1.34, 95% CI: 1.11 to 1.63), Pakistani (ORs: 1.79, 95% CI: 1.42 to 2.27), Any other Asian background (ORs: 1.45, 95% CI: 1.07 to 1.95), African (ORs: 1.30, 95% CI: 1.03 to 1.63), Caribbean (ORs: 1.79, 95% CI: 1.31 to 2.43), Mixed White and Black Caribbean (ORs: 2.65, 95% CI: 1.88 to 3.71), Any other mixed/multiple background (ORs: 2.01, 95% CI: 1.45 to 2.71), Eastern European (ORs: 1.92, 95% CI: 1.36 to 2.72), Any other White background (ORs: 1.28, 95% CI: 1.03 to 1.58), and Any other ethnic group background (ORs: 2.03, 95% CI: 1.37 to 3.00).

**Table 2.** Weight adjusted ethnic differences in loneliness across 21 ethnic groups in different models (N=13967).

| Ethnicity | Model 1 | Model 2 | Model 3 | Model 4 |
| --- | --- | --- | --- | --- |
|  | (without adjusted for any covariate) | (adjusted on individual level <sup>a</sup>) | (adjusted on individual, and social and household levels <sup>b</sup>) | (adjusted on individual, social and household, and socio-economic and environmental levels <sup>c</sup>) |
|  | Odds ratio (95% CI) | Odds ratio (95% CI) | Odds ratio (95% CI) | Odds ratio (95% CI) |
| White British (Reference) | 1.00 | 1.00 | 1.00 | 1.00 |
| Asian: Bangladeshi | 1.46 (1.03 to 2.06) * | 1.08 (0.76 to 1.55) | 1.22 (0.85 to 1.75) | 0.87 (0.59 – 1.30) |
| Asian: Chinese | 1.29 (0.97 to 1.73) | 0.98 (0.71 to 1.33) | 1.07 (0.78 to 1.47) | 0.93 (0.66 – 1.32) |
| Asian: Indian | 1.34 (1.11 to 1.63) ** | 1.14 (0.94 to 1.40) | 1.38 (1.13 to 1.70) ** | 1.21 (0.97 – 1.50) |
| Asian: Pakistani | 1.79 (1.42 to 2.27) *** | 1.43 (1.12 to 1.84) ** | 1.69 (1.31 to 2.17) *** | 1.33 (1.01 – 1.73) * |
| Any other Asian background | 1.45 (1.07 to 1.95) * | 1.18 (0.88 to 1.57) | 1.36 (0.99 to 1.88) | 1.14 (0.80 – 1.62) |
| Black: African | 1.30 (1.03 to 1.63) * | 1.00 (0.78 to 1.29) | 1.12 (0.87 to 1.44) | 0.93 (0.70 – 1.23) |
| Black: Caribbean | 1.79 (1.31 to 2.43) *** | 1.61 (1.15 to 2.23) ** | 1.55 (1.12 to 2.15) ** | 1.37 (0.97 – 1.94) |
| Any other Black/African/Caribbean background | 1.60 (0.98 to 2.60) | 1.25 (0.78 to 2.01) | 1.24 (0.73 to 2.13) | 1.08 (0.65 – 1.81) |
| Mixed: White and Asian | 1.33 (0.99 to 1.79) | 0.96 (0.71 to 1.31) | 1.01 (0.75 to 1.38) | 0.86 (0.63 – 1.20) |
| Mixed: White and Black African | 1.69 (1.00 to 2.86) | 1.29 (0.74 to 2.24) | 1.19 (0.72 to 1.99) | 1.08 (0.65 – 1.82) |
| Mixed: White and Black Caribbean | 2.65 (1.88 to 3.71) *** | 1.89 (1.34 to 2.67) *** | 1.82 (1.27 to 2.59) ** | 1.77 (1.20 – 2.62) ** |
| Any other mixed/multiple background | 2.01 (1.45 to 2.78) *** | 1.50 (1.06 to 2.11) * | 1.55 (1.06 to 2.27) * | 1.29 (0.86 – 1.94) |
| White: Eastern European | 1.92 (1.36 to 2.72) *** | 1.52 (1.03 to 2.24) * | 1.69 (1.14 to 2.52) ** | 1.60 (1.04 – 2.46) * |
| White: Gypsy/Traveller | 0.97 (0.54 to 1.73) | 0.76 (0.44 to 1.32) | 0.85 (0.51 to 1.42) | 0.75 (0.44 – 1.26) |
| White: Irish | 1.00 (0.63 to 1.60) | 1.01 (0.62 to 1.65) | 1.04 (0.64 to 1.67) | 0.95 (0.60 – 1.50) |
| White: Roma | 0.66 (0.34 to 1.29) | 0.49 (0.25 to 0.97) * | 0.43 (0.21 to 0.86) * | 0.39 (0.20 – 0.77) ** |
| Any other White background | 1.28 (1.03 to 1.58) * | 1.02 (0.81 to 1.28) | 1.03 (0.81 to 1.30) | 0.95 (0.74 – 1.23) |
| Other: Arab | 1.60 (0.94 to 2.70) | 1.23 (0.69 to 2.17) | 1.40 (0.77 to 2.56) | 1.08 (0.62 – 1.90) |
| Any other ethnic group | 2.03 (1.37 to 3.00) *** | 1.75 (1.14 to 2.69) * | 1.85 (1.22 to 2.83) ** | 1.39 (0.89 – 2.16) |
| Jewish | 0.93 (0.74 to 1.17) | 0.95 (0.75 to 1.20) | 1.08 (0.85 to 1.39) | 0.98 (0.74 – 1.31) |

\* P < 0.05; \*\* P < 0.01; \*\*\* P < 0.001. <sup>a</sup> : adjusted for age, sex, and previous COVID infection; <sup>b</sup> : adjusted for age, sex, previous COVID infection, marital/partnership status, and household generational structure; <sup>c</sup> : adjusted for age, sex, previous COVID infection, marital/partnership status, household generational structure, financial concerns, region of residence, education, and employment status.

After the adjustments in Models 2, 3, and 4, fewer ethnic groups showed significantly higher odds compared to the White British group. Adjusting for individual factors (Model 2) eliminated significant differences for groups including Bangladeshi, Indian, African, and Any other White background, suggesting age and sex largely explained the differences due to the insignificant effect of previous COVID infection. In Model 4, adjusting for all socio-demographic factors eliminated significant differences for the Caribbean, Any other Mixed/Multiple background, and Any other ethnic groups. Pakistani (Model 4 ORs: 1.33, 95% CI: 1.01 to 1.73), Mixed White and Black Caribbean (Model 4 ORs: 1.77, 95% CI: 1.20 to 2.62), and Eastern European (Model 4 ORs: 1.60, 95% CI: 1.04 to 2.46) had significantly higher odds of loneliness in all 4 models.

Only the Roma group had significantly lower odds of loneliness than the White British group, but this was only observed in the adjusted models (Model 2 OR: 0.49, 95% CI: 0.25 to 0.97; Model 3 OR: 0.43, 95% CI: 0.21 to 0.86; Model 4 OR: 0.39, 95% CI: 0.20 to 0.77). The complete results of the weighted logistic regression analyses are shown in Table S2. The sensitivity analysis using the continuous loneliness score (Table S3) and the complete case analysis (Table S4) all showed a similar pattern of results.

### Ethnic inequalities in loneliness across 6 aggregated ethnic groups

Compared to White British (Table 3), all other aggregated ethnic groups had significantly higher odds of loneliness in the unadjusted model (Model 1), including White minority (ORs: 1.39, 95% CI: 1.16 to 1.65), Mixed or multiple ethnic groups (ORs: 1.92, 95% CI: 1.59 to 2.32), Asian (ORs: 1.47, 95% CI: 1.28 to 1.69), Black (ORs: 1.46, 95% CI: 1.21 to 1.75), and Other ethnic groups (ORs: 1.68, 95% CI: 1.23 to 2.28). Most of those ethnic minority groups continued to have significantly higher odds of loneliness in adjusted models 2 and 3. Only Mixed or multiple ethnic groups showed significantly higher odds of loneliness (ORs: 1.27, 95% CI: 1.01 to 1.58) compared to White British after adjusting for all factors in Model 4. The complete results of the weighted logistic regression analyses using six aggregated ethnic groups are presented in Table S5.

**Table 3.** Weight adjusted ethnic differences in loneliness across 6 ethnic groups in different models (N=13967).

| Ethnicity | Model 1 | Model 2 | Model 3 | Model 4 |
| --- | --- | --- | --- | --- |
|  | (without adjusted for any covariate) | (adjusted on individual level) | (adjusted on individual, and social and household levels) | (adjusted on individual, social and household, and socio-economic and environmental levels) |
|  | Odds ratio (95% CI) | Odds ratio (95% CI) | Odds ratio (95% CI) | Odds ratio (95% CI) |
| White British (Reference) | 1.00 | 1.00 | 1.00 | 1.00 |
| White minority | 1.39 (1.16 to 1.65) *** | 1.13 (0.94 to 1.37) | 1.18 (0.97 to 1.44) | 1.10 (0.89 – 1.36) |
| Mixed or multiple ethnic groups | 1.92 (1.59 to 2.32) *** | 1.41 (1.15 to 1.73) ** | 1.41 (1.14 to 1.74) ** | 1.27 (1.01 – 1.58) * |
| Asian | 1.47 (1.28 to 1.69) *** | 1.19 (1.03 to 1.38) * | 1.39 (1.19 to 1.62) *** | 1.16 (0.98 – 1.37) |
| Black | 1.46 (1.21 to 1.75) *** | 1.18 (0.96 to 1.44) | 1.25 (1.02 to 1.52) * | 1.07 (0.85 – 1.34) |
| Other ethnic groups | 1.68 (1.23 to 2.28) ** | 1.51 (1.08 to 2.11) * | 1.63 (1.17 to 2.27) ** | 1.29 (0.91 – 1.83) |

\* P < 0.05; \*\* P < 0.01; \*\*\* P < 0.001.

## Discussion

### Summary

This study provides the most comprehensive analysis of ethnic inequalities in loneliness in Britain, examining 21 disaggregated and 6 aggregated ethnic groups during the COVID-19 pandemic. Without accounting for other factors, over half of the disaggregated ethnic minority groups showed significantly higher levels of loneliness compared to White British, while the aggregated analysis suggested higher loneliness across all minority groups. The disaggregated analysis revealed nuanced ethnic disparities in loneliness that were largely overlooked by the aggregated analysis. After controlling for socio-demographic differences, Pakistani, Mixed White and Black Caribbean, and Eastern European groups had higher levels of loneliness, while Roma had lower levels of loneliness compared to White British. Aggregated analyses only identified higher loneliness in the Mixed/Multiple ethnic groups.

### Comparison to previous literature and interpretation of findings

This study found clear evidence of ethnic disparities, with the disaggregated approach revealing more nuanced differences than the aggregated analysis. Without adjusting for covariates, the aggregated analysis suggested that all ethnic minorities had higher odds of loneliness compared to White British, overlooking variations among subgroups. For instance, within the Asian group, Bangladeshi, Indian, Pakistani, and those of Any other Asian background had higher odds of loneliness, while no difference was observed between Chinese and White British. After adjusting for covariates, the aggregated analysis found higher odds of loneliness only among Mixed/Multiple ethnic groups, while the disaggregated analysis also identified higher loneliness among Pakistani and Eastern European people, and lower loneliness among Roma. These nuanced ethnic differences, highlighting unique challenges faced by specific groups—such as labor market discrimination, especially during the pandemic (Fancourt, Steptoe, and Bradbury 2022)— can only be captured through more granular approaches and the data on underrepresented groups (Irizar et al. 2024). Additionally, in aggregated analyses, the increased loneliness in some subgroups and reduced loneliness in others were centralized, resulting in masked variations. This may also explain the absence of ethnic differences in loneliness in previous studies using aggregated methods (O’Connor et al. 2021).

Three distinct patterns in ethnic inequalities in loneliness emerged when comparing covariate-unadjusted and adjusted models. First, controlling for covariates shifted some differences between White British and several ethnic minority groups from significant to insignificant. For example, after adjusting for individual factors, disparities between White British and groups such as Bangladeshi, Indian, and African disappeared. As age and sex were the most significant factors in this model, it suggests that differences in population structure largely explained these differences. Additionally, after adjusting for socio-economic and environmental factors, differences between British White and groups like Indian, Caribbean, and Any other mixed/multiple background also became insignificant. These findings align with prior research identifying individual and socio-economic factors as key drivers of loneliness (Barjaková, Garnero, and d’Hombres 2023; McQuaid et al. 2021). These findings highlight the need for targeted support (Salway et al. 2020), such as interventions to reduce loneliness among youth in certain ethnic minority background (Lenoir and Wong 2023).

The second pattern contrasted with the above one. After covariate adjustment, some ethnic groups exhibited significant differences in loneliness compared to the White British group, which were previously insignificant. For instance, introducing social and household-level covariates slightly increased disparities in several groups, leading to significant differences in the Indian group. Additionally, the Roma group showed significantly lower odds of loneliness only after these covariates were included. This pattern also appeared in the Black group in aggregated analyses. These findings suggested that certain covariates acted as suppressor variables (Ludlow and Klein 2014), revealing some ethnic differences in loneliness that might not be evident without their inclusion, meanwhile largely explaining ethnic differences in other groups. Thus, it would not be surprising that the same intervention might reduce loneliness in some groups while increasing it in others.

The final pattern revealed that, after covariate adjustment, some ethnic groups not only had reduced differences in loneliness compared to the White British group, but also had lower odds of loneliness, despite previously showing higher odds—though none of these reversed differences were statistically significant. This was observed in groups such as Bangladeshi, Chinese, African, Mixed White and Asian, Irish, and Any other White background after adjusting for socio-demographic factors. This pattern aligns with previous findings showing reversed inequalities in loneliness once these factors were accounted for (Fokkema and Naderi 2013; Visser and El Fakiri 2016). This indicated that some ethnic groups might be more sensitive to specific targeted interventions for reducing loneliness during the crisis than others. Rather than applying a “one-size-fits-all” approach, policymakers should consider the effectiveness of certain interventions on different ethnic communities.

### Strengths and limitations

This study is a secondary analysis of the EVENS dataset, which includes one of the largest and most diverse samples of ethnic minority individuals in Britain during the pandemic. The large sample size, particularly for underrepresented populations, allowed for a comprehensive analysis across a wide range of ethnic groups. However, the EVENS dataset was collected between February and November 2021, missing the early period of the pandemic, particularly the first lockdown, although many studies have shown the pandemic’s persistent and prolonged impact on mental health (Manchia et al. 2022).

Additionally, due to the many statistical tests conducted in this study, there were increased risks of type I errors (Bender and Lange 2001). Due to the exploratory nature of this study, the choice of not applying multiple comparison corrections in this study reflected the preference for pursuing possible discoveries (Rothman 1990), even if they require future verification.

## Conclusions

This study provided clear evidence of ethnic inequalities in loneliness during COVID-19 pandemic in Britain, indicating higher levels of loneliness in more than half groups of ethnic minorities in Britain. After accounting for various socio-demographic factors, significantly higher levels of loneliness were found in Pakistani, Mixed White and Black Caribbean, and Eastern European groups, and significantly lower levels of loneliness were found in the Roma group compared to the White British group. These findings underscore the need for targeted support for specific ethnic minority groups. As we move beyond the pandemic, further research is necessary to determine whether these disparities persist and to guide the development of interventions that effectively address the unique needs of each group.

## Data Availability

All data produced are available online at https://doi.org/10.5255/UKDA-SN-9116-1.

## Acknowledgments

We are grateful to the EVENS participants and the EVENS team for their invaluable contributions. We would also like to thank the members of the public who provided valuable insights during the initial stage of this research, helping to inform our analysis plan. We extend our special thanks to Harry Taylor for his valuable feedback on the manuscript.

