## Supplemental material for "Ethnic inequalities in loneliness in Britain during the COVID-19 pandemic: Evidence for Equality National Survey (EVENS)"

### Methods

#### Outcome variable

The UCLA 3-item Loneliness Scale included questions of “How often do you feel you lack companionship?”, “How often do you feel left out?”, and “How often do you feel isolated from others?”. Response options for each question were 1 "Hardly ever or never", 2 "Some of the time", or 3 "Often". All items were summed up arithmetically to give a total score of loneliness ranging from 3 to 9, with higher scores indicating more evidence of loneliness.

#### Covariates

Detailed categories for covariates included in this study are listed below:

- Age: The age range in the EVENS dataset included individuals aged 18 years and older, aligning with the inclusion criteria. Categorized age groups were defined as 18-24, 25-34, 35-44, 45-54, 55-64, and 65-75 years old.
- Sex: Sex (registered at birth) was defined as female and male.
- Previous COVID infections: Whether participants ever had any kind of test for coronavirus (Covid-19) and whether they ever received a positive result for such tests were asked. Four categories were created for analysis including 1 “Ever had positive results”, 2 “Never tested”, 3 “Never had positive results”, 4 “Other/Prefer not to say”.
- Marital/Partnership status: The original marital/partnership status in the EVENS dataset comprised of 10 categories, including 1 “Never married and never registered a civil partnership”, 2 “Married”, 3 “In a registered civil partnership”, 4 “Separated, but still legally married”, 5 “Separated, but still legally in a civil partnership”, 6 “Divorced”, 7 “Formerly in a civil partnership which is now legally dissolved”, 8 “Widowed”, 9 “Surviving partner from a registered civil partnership”, 10 “Prefer not to say”. The 10 statuses were then categorized into 5 statuses for analysis including 1 "Married/Civil", 2 "Not married/not civil", 3 "Separated/ Divorced/Dissolved", 4 "Widowed/Partner died", and 5 “prefer not to say”.

- Number of generations living in the household: The EVENS study asked the question of “How many generations live in your current household?” with 4 categories of 1 “One generation”, 2 “Two generations”, 3 “Three generations”, and 4 “Other/Prefer not to say” for analysis in this study.
- Worrying about financial situation: The EVENS study asked the question of “How worried, if at all, are you about your future financial situation?” with 5 options of 1 “Not at all worried”, 2 “Somewhat worried”, 3 “Very worried”, 4 “Extremely worried”, and 5 “Prefer not to say”. The 5 categories were used for analysis in the current study.
- Region: The original region variable in the EVENS dataset comprised of 11 regions, including 1 “North East”, 2 “North West”, 3 “Yorkshire and Humber”, 4 “West Midlands”, 5 “East Midlands”, 6 “East of England”, 7 “South West”, 8 “South East”, 9 “London”, 10 “Wales”, 11 “Scotland”. The 11 regions were then categorized into 4 main regions for analysis including 1 "North + Scotland", 2 "Midland + Wales", 3 "South", and 4 "London".
- Education: The highest educational or professional qualification of the participant was asked with seven options in the EVENS. The categories were aggregated for analysis with four final categories, including 1 “Degree or equivalent” (University higher degree/ First degree level qualification), 2 “Higher education” (Diplomas in higher education, HNC/HND/BTEC Higher or equivalent), 3 “GCE, A level, GCSE or equivalent” (A-Level, Scottish Higher, Welsh Baccalaureate, International Baccalaureate or equivalent/ Vocational qualification/GCSE/O-Level/CSE), and 4 “Other/no qualification”.
- Employment: The current employment situations of participants were asked with fourteen options in the EVENS. The categories were recoded for analysis with four final categories, including 1 “Employed” (Self-employed/ In full-time paid employment/ In part-time paid employment/ On a government training scheme/ Unpaid worker in family business/ Working in an apprenticeship), 2 “Not in labor force” (Retired/ On maternity leave/ Looking after family or home/ Full-time

student/ Long-term sick or disabled), 3 “Unemployed”, and 4 “Other/Prefer not to say”.

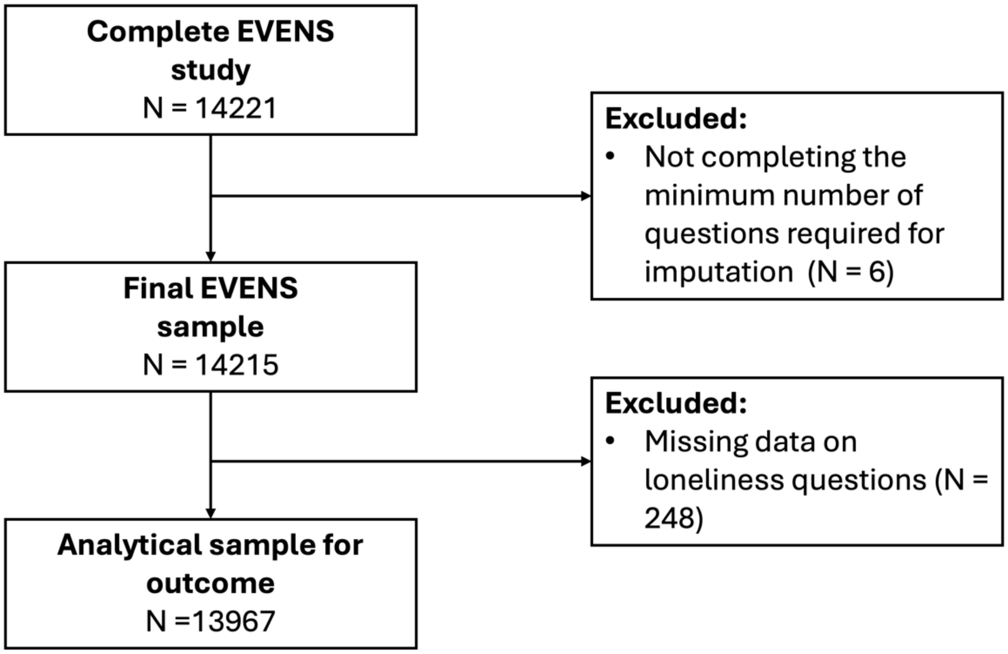


Figure S1. Participant flow diagram.

**Statistical analysis**

To check multicollinearity among the variables, the variance inflation factor (VIF) was assessed, with values less than 10 considered acceptable [1]. There was no evidence of multicollinearity as VIF scores were less than 10 (VIF mean = 1.40, VIF Min = 1.03, VIF Max = 3.07) (Table S6). The linearity of logit assumption was not checked because there was no continuous variable in the model.

Table S1. Sample characteristics for other variables (Unweighted Ns and weighted %, N = 14215).

| **Variable (Continuous)** | **Unweighted frequency N** | **Weighted percentage %** |
|---|---|---|
| **Previous COVID infection** | | |
| Ever had positive results | 1436 | 6.94 |
| Never tested | 4698 | 40.32 |
| Never had positive results | 7751 | 50.95 |
| Other/Prefer not to say | 330 | 1.79 |
| **Marital/Partnership status** | | |
| Married/Civil | 6488 | 47.13 |
| Not married/not civil | 5889 | 35.35 |
| Separated/Divorced/Dissolved | 1173 | 10.05 |
| Widowed/Partner died | 310 | 5.58 |
| Prefer not to say | 355 | 1.89 |
| **Number of generations living in household** | | |
| One generation | 6465 | 56.15 |
| Two generations | 7022 | 40.65 |
| Three generations | 525 | 2.26 |
| Other/Prefer not to say | 203 | 0.94 |
| **Worrying about financial situation** | | |
| Not at all worried | 4077 | 38.85 |
| Somewhat worried | 6964 | 44.79 |
| Very worried | 1659 | 8.53 |
| Extremely worried | 1140 | 5.60 |
| Prefer not to say | 375 | 2.23 |
| **Region of residence** | | |
| North + Scotland | 3864 | 32.42 |
| Midland + Wales | 4360 | 31.23 |
| South | 2417 | 22.97 |
| London | 3574 | 13.38 |
| **Education** | | |
| Degree or equivalent | 7837 | 36.78 |
| Higher education | 1423 | 8.68 |
| GCE, A level, GCSE or equivalent | 4201 | 48.41 |
| Other/no qualification | 754 | 6.12 |
| **Employment** | | |
| Employed | 9192 | 62.32 |
| Not in labor force | 3840 | 32.79 |
| Unemployed | 904 | 3.64 |
| Other/Prefer not to say | 279 | 1.25 |
| **Total** | 14215 | 100 |

Table S2. Weight adjusted differences in loneliness across different models using 21 disaggregated ethnic groups (N=13967).

| Variable | Model 1 (without adjusted for any covariate) | Model 2 (adjusted on individual level [a]) | Model 3 (adjusted on individual, and social and household levels [b]) | Model 4 (adjusted on individual, social and household, and socio-economic and environmental levels [c]) |
|---|---|---|---|---|
| | Odds ratio (95% CI) | Odds ratio (95% CI) | Odds ratio (95% CI) | Odds ratio (95% CI) |
| **Ethnicity** | | | | |
| White British (Reference) | 1.00 | 1.00 | 1.00 | 1.00 |
| Asian: Bangladeshi | 1.46 (1.03 to 2.06) * | 1.08 (0.76 to 1.55) | 1.22 (0.85 to 1.75) | 0.87 (0.59 to 1.30) |
| Asian: Chinese | 1.29 (0.97 to 1.73) | 0.98 (0.71 to 1.33) | 1.07 (0.78 to 1.47) | 0.93 (0.66 to 1.32) |
| Asian: Indian | 1.34 (1.11 to 1.63) ** | 1.14 (0.94 to 1.40) | 1.38 (1.13 to 1.70) ** | 1.21 (0.97 to 1.50) |
| Asian: Pakistani | 1.79 (1.42 to 2.27) *** | 1.43 (1.12 to 1.84) ** | 1.69 (1.31 to 2.17) *** | 1.33 (1.01 to 1.73) * |
| Any other Asian background | 1.45 (1.07 to 1.95) * | 1.18 (0.88 to 1.57) | 1.36 (0.99 to 1.88) | 1.14 (0.80 to 1.62) |
| Black: African | 1.30 (1.03 to 1.63) * | 1.00 (0.78 to 1.29) | 1.12 (0.87 to 1.44) | 0.93 (0.70 to 1.23) |
| Black: Caribbean | 1.79 (1.31 to 2.43) *** | 1.61 (1.15 to 2.23) ** | 1.55 (1.12 to 2.15) ** | 1.37 (0.97 to 1.94) |
| Any other Black/African/Caribbean background | 1.60 (0.98 to 2.60) | 1.25 (0.78 to 2.01) | 1.24 (0.73 to 2.13) | 1.08 (0.65 to 1.81) |
| Mixed: White and Asian | 1.33 (0.99 to 1.79) | 0.96 (0.71 to 1.31) | 1.01 (0.75 to 1.38) | 0.86 (0.63 to 1.20) |
| Mixed: White and Black African | 1.69 (1.00 to 2.86) | 1.29 (0.74 to 2.24) | 1.19 (0.72 to 1.99) | 1.08 (0.65 to 1.82) |
| Mixed: White and Black Caribbean | 2.65 (1.88 to 3.71) *** | 1.89 (1.34 to 2.67) *** | 1.82 (1.27 to 2.59) ** | 1.77 (1.20 to 2.62) ** |
| Any other mixed/multiple background | 2.01 (1.45 to 2.78) *** | 1.50 (1.06 to 2.11) * | 1.55 (1.06 to 2.27) * | 1.29 (0.86 to 1.94) |
| White: Eastern European | 1.92 (1.36 to 2.72) *** | 1.52 (1.03 to 2.24) * | 1.69 (1.14 to 2.52) ** | 1.60 (1.04 to 2.46) * |
| White: Gypsy/Traveller | 0.97 (0.54 to 1.73) | 0.76 (0.44 to 1.32) | 0.85 (0.51 to 1.42) | 0.75 (0.44 to 1.26) |
| White: Irish | 1.00 (0.63 to 1.60) | 1.01 (0.62 to 1.65) | 1.04 (0.64 to 1.67) | 0.95 (0.60 to 1.50) |
| White: Roma | 0.66 (0.34 to 1.29) | 0.49 (0.25 to 0.97) * | 0.43 (0.21 to 0.86) * | 0.39 (0.20 to 0.77) ** |
| Any other White background | 1.28 (1.03 to 1.58) * | 1.02 (0.81 to 1.28) | 1.03 (0.81 to 1.30) | 0.95 (0.74 to 1.23) |
| Other: Arab | 1.60 (0.94 to 2.70) | 1.23 (0.69 to 2.17) | 1.40 (0.77 to 2.56) | 1.08 (0.62 to 1.90) |
| Any other ethnic group | 2.03 (1.37 to 3.00) *** | 1.75 (1.14 to 2.69) * | 1.85 (1.22 to 2.83) ** | 1.39 (0.89 to 2.16) |
| Jewish | 0.93 (0.74 to 1.17) | 0.95 (0.75 to 1.20) | 1.08 (0.85 to 1.39) | 0.98 (0.74 to 1.31) |

| | | | | |
|---|---|---|---|---|
| **Age (years old)** | - | | | |
| 18-24 (Reference) | - | 1.00 | 1.00 | 1.00 |
| 25-34 | - | 0.71 (0.53 to 0.94) * | 0.83 (0.62 to 1.12) | 0.77 (0.57 to 1.03) |
| 35-44 | - | 0.62 (0.47 to 0.82) ** | 0.80 (0.60 to 1.08) | 0.74 (0.55 to 1.01) |
| 45-54 | - | 0.50 (0.38 to 0.66) *** | 0.64 (0.47 to 0.86) ** | 0.57 (0.41 to 0.77) *** |
| 55-64 | - | 0.43 (0.32 to 0.57) *** | 0.57 (0.41 to 0.79) ** | 0.53 (0.38 to 0.73) *** |
| 65-75 | | 0.26 (0.20 to 0.35) *** | 0.31 (0.22 to 0.45) *** | 0.32 (0.23 to 0.47) *** |
| **Sex** | - | | | |
| Male (Reference) | - | 1.00 | 1.00 | 1.00 |
| Female | | 1.34 (1.17 to 1.52) *** | 1.26 (1.10 to 1.45) ** | 1.14 (0.99 to 1.32) |
| **Previous COVID infection** | - | | | |
| Ever had positive results (Reference) | - | 1.00 | 1.00 | 1.00 |
| Never tested | - | 0.95 (0.73 to 1.24) | 0.90 (0.68 to 1.18) | 0.88 (0.67 to 1.17) |
| Never had positive results | - | 0.97 (0.75 to 1.26) | 0.92 (0.71 to 1.20) | 0.89 (0.68 to 1.17) |
| Other/Prefer not to say | | 1.49 (0.90 to 2.48) | 1.38 (0.85 to 2.25) | 1.14 (0.68 to 1.90) |
| **Marital/Partnership status** | - | | | |
| Married/Civil (Reference) | - | - | 1.00 | 1.00 |
| Not married/not civil | - | - | 2.00 (1.68 to 2.38) *** | 1.83 (1.53 to 2.18) *** |
| Separated/Divorced/Dissolved | - | - | 2.55 (2.04 to 3.17) *** | 2.34 (1.85 to 2.97) *** |
| Widowed/Partner died | - | - | 3.27 (2.29 to 4.67) *** | 3.12 (2.14 to 4.55) *** |
| Prefer not to say | | - | 1.83 (1.08 to 3.08) * | 1.57 (0.91 to 2.70) |
| **Number of generations living in household** | - | | | |
| One generation (Reference) | - | - | 1.00 | 1.00 |
| Two generations | - | - | 0.99 (0.85 to 1.15) | 0.92 (0.78 to 1.08) |
| Three generations | - | - | 1.14 (0.78 to 1.67) | 1.00 (0.69 to 1.46) |
| Other/Prefer not to say | | - | 1.35 (0.72 to 2.54) | 1.21 (0.63 to 1.32) |
| **Worrying about financial situation** | - | | | |
| Not at all worried (Reference) | - | - | - | 1.00 |
| Somewhat worried | - | - | - | 2.66 (2.24 to 3.15) *** |
| Very worried | - | - | - | 4.74 (3.62 to 6.21) *** |
| Extremely worried | - | - | - | 7.46 (5.55 to 10.02) *** |

| | | | | |
|---|---|---|---|---|
| Prefer not to say | | - | - | 2.64 (1.66 to 4.20) *** |
| **Region of residence** | - | | | |
| North + Scotland (Reference) | - | - | - | 1.00 |
| Midland + Wales | - | - | - | 0.91 (0.76 to 1.09) |
| South | - | - | - | 0.96 (0.79 to 1.18) |
| London | | - | - | 0.99 (0.81 to 1.21) |
| **Education** | - | | | |
| Degree or equivalent (Reference) | - | - | - | 1.00 |
| Higher education | - | - | - | 0.94 (0.81 to 1.09) |
| GCE, A level, GCSE or equivalent | - | - | - | 0.82 (0.58 to 1.17) |
| Other/no qualification | | - | - | 0.69 (0.43 to 1.08) |
| **Employment** | - | | | |
| Employed (Reference) | - | - | - | 1.00 |
| Not in labor force | - | - | - | 1.39 (1.14 to 1.69) ** |
| Unemployed | - | - | - | 1.35 (1.04 to 1.76) |
| Other/Prefer not to say | | - | - | 1.42 (0.83 to 2.43) |

* P < 0.05; ** P < 0.01; *** P < 0.001. [a] : adjusted for age, sex, and previous COVID infection; [b] : adjusted for age, sex, previous COVID infection, marital/partnership status, and household generational structure; [c] : adjusted for age, sex, previous COVID infection, marital/partnership status, household generational structure, financial concerns, region of residence, education, and employment status.

Table S3. Sensitivity analysis: weighted linear regression analysis with continuous loneliness scores using 21 disaggregated ethnic groups (N=13967).

| **Variable** | **Model 1** (without adjusted for any covariate) | **Model 2** (adjusted on individual level[a]) | **Model 3** (adjusted on individual, and social and household levels[b]) | **Model 4** (adjusted on individual, social and household, and socio-economic and environmental levels[c]) |
|---|---|---|---|---|
| | Coefficient (95% CI) | Coefficient (95% CI) | Coefficient (95% CI) | Coefficient (95% CI) |
| **Ethnicity** | | | | |
| White British (Reference) | 0.00 | 0.00 | 0.00 | 0.00 |
| Asian: Bangladeshi | 0.46 (0.16 to 0.76) ** | 0.15 (-0.16 to 0.46) | 0.26 (-0.04 to 0.56) | -0.06 (-0.36 to 0.25) |
| Asian: Chinese | 0.25 (-0.02 to 0.51) | -0.05 (-0.33 to 0.22) | 0.03 (-0.24 to 0.31) | -0.07 (-0.34 to 0.20) |
| Asian: Indian | 0.34 (0.15 to 0.52) *** | 0.17 (-0.16 to 0.35) | 0.34 (0.16 to 0.52) *** | 0.20 (0.02 to 0.37) * |
| Asian: Pakistani | 0.58 (0.33 to 0.82) *** | 0.32 (0.08 to 0.56) * | 0.45 (0.22 to 0.68) *** | 0.18 (-0.05 to 0.41) |
| Any other Asian background | 0.30 (0.06 to 0.55) * | 0.08 (-0.16 to 0.32) | 0.21 (-0.03 to 0.46) | 0.06 (-0.19 to 0.31) |
| Black: African | 0.26 (0.04 to 0.49) * | -0.01 (-0.25 to 0.23) | 0.11 (-0.12 to 0.33) | -0.07 (-0.30 to 0.17) |
| Black: Caribbean | 0.68 (0.36 to 1.00) *** | 0.55 (0.22 to 0.88) ** | 0.50 (0.19 to 0.82) ** | 0.37 (0.08 to 0.65) * |
| Any other Black/African/Caribbean background | 0.41 (0.03 to 0.80) * | 0.13 (-0.22 to 0.49) | 0.12 (-0.29 to 0.52) | -0.02 (-0.36 to 0.33) |
| Mixed: White and Asian | 0.34 (0.08 to 0.60) * | -0.01 (-0.27 to 0.25) | 0.03 (-0.21 to 0.28) | -0.10 (-0.32 to 0.13) |
| Mixed: White and Black African | 0.42 (-0.01 to 0.85) | 0.12 (-0.29 to 0.53) | 0.04 (-0.34 to 0.42) | -0.05 (-0.43 to 0.32) |
| Mixed: White and Black Caribbean | 1.04 (0.69 to 1.40) *** | 0.66 (0.31 to 1.00) *** | 0.61 (0.25 to 0.96) ** | 0.54 (0.18 to 0.89) ** |
| Any other mixed/multiple background | 0.75 (0.46 to 1.03) *** | 0.41 (0.12 to 0.69) ** | 0.43 (0.14 to 0.73) ** | 0.24 (-0.04 to 0.52) |
| White: Eastern European | 0.59 (0.26 to 0.92) *** | 0.33 (-0.02 to 0.68) | 0.42 (0.07 to 0.77) * | 0.35 (-0.004 to 0.70) |
| White: Gypsy/Traveller | -0.03 (-0.49 to 0.43) | -0.28 (-0.69 to 0.13) | -0.15 (-0.54 to 0.23) | -0.30 (-0.69 to 0.08) |
| White: Irish | -0.06 (-0.43 to 0.31) | -0.05 (-0.41 to 0.30) | -0.03 (-0.36 to 0.31) | -0.09 (-0.40 to 0.21) |
| White: Roma | -0.39 (-0.84 to 0.06) | -0.70 (-1.15 to -0.26) ** | -0.86 (-1.31 to -0.41) *** | -0.93 (-1.37 to -0.50) *** |
| Any other White background | 0.29 (0.08 to 0.50) ** | 0.05 (-0.16 to 0.26) | 0.06 (-0.15 to 0.27) | 0.01 (-0.20 to 0.21) |
| Other: Arab | 0.59 (0.12 to 1.06) * | 0.30 (-0.23 to 0.84) | 0.42 (-0.14 to 0.99) | 0.16 (-0.35 to 0.67) |
| Any other ethnic group | 0.60 (0.30 to 0.89) *** | 0.42 (0.11 to 0.74) ** | 0.46 (0.15 to 0.77) ** | 0.16 (0.13 to 0.46) |
| Jewish | -0.02 (-0.23 to 0.20) | -0.002 (-0.22 to 0.21) | 0.13 (-0.09 to 0.34) | 0.08 (-0.15 to 0.30) |

| | | | | |
|---|---|---|---|---|
| **Age (years old)** | - | | | |
| 18-24 (Reference) | - | 0.00 | 0.00 | 0.00 |
| 25-34 | - | -0.36 (-0.61 to -0.10) ** | -0.19 (-0.45 to 0.07) | -0.24 (-0.48 to -0.002) * |
| 35-44 | - | -0.54 (-0.81 to -0.28) *** | -0.26 (-0.53 to 0.02) | -0.30 (-0.56 to -0.05) * |
| 45-54 | - | -0.80 (-1.05 to -0.54) *** | -0.51 (-0.78 to -0.25) *** | -0.58 (-0.83 to -0.34) *** |
| 55-64 | - | -0.93 (-1.20 to -0.66) *** | -0.60 (-0.89 to -0.31) *** | -0.61 (-0.88 to -0.35) *** |
| 65-75 | | -1.35 (-1.60 to -1.10) *** | -1.08 (-1.37 to -0.78) *** | -0.99 (-1.27 to -0.72) *** |
| **Sex** | - | | | |
| Male (Reference) | - | 0.00 | 0.00 | 0.00 |
| Female | | 0.36 (0.24 to 0.47) *** | 0.30 (0.18 to 0.41) *** | 0.19 (0.08 to 0.30) ** |
| **Previous COVID infection** | - | | | |
| Ever had positive results (Reference) | - | 0.00 | 0.00 | 0.00 |
| Never tested | - | -0.05 (-0.29 to 0.19) | -0.11 (-0.35 to 0.13) | -0.13 (-0.35 to 0.09) |
| Never had positive results | - | -0.04 (-0.27 to 0.19) | -0.09 (-0.32 to 0.14) | -0.11 (-0.33 to 0.10) |
| Other/Prefer not to say | | 0.51 (0.04 to 0.98) * | 0.42 (0.03 to 0.87) | 0.20 (-0.20 to 0.59) |
| **Marital/Partnership status** | - | | | |
| Married/Civil (Reference) | - | - | 0.00 | 0.00 |
| Not married/not civil | - | - | 0.73 (0.58 to 0.88) *** | 0.60 (0.45 to 0.73) *** |
| Separated/Divorced/Dissolved | - | - | 0.89 (0.70 to 1.09) *** | 0.75 (0.56 to 0.94) *** |
| Widowed/Partner died | - | - | 1.07 (0.79 to 1.35) *** | 0.95 (0.69 to 1.20) *** |
| Prefer not to say | | - | 0.41 (-0.04 to 0.86) | 0.24 (-0.20 to 0.68) |
| **Number of generations living in household** | - | | | |
| One generation (Reference) | - | - | 0.00 | 0.00 |
| Two generations | - | - | 0.42 (-0.14 to 0.99) | -0.11 (-0.24 to 0.01) |
| Three generations | - | - | 0.46 (0.15 to 0.77) | 0.23 (-0.12 to 0.57) |
| Other/Prefer not to say | | - | 0.13 (-0.09 to 0.34) | 0.30 (-0.32 to 0.91) |
| **Worrying about financial situation** | - | | | |
| Not at all worried (Reference) | - | - | - | 0.00 |
| Somewhat worried | - | - | - | 0.85 (0.73 to 0.97) *** |
| Very worried | - | - | - | 1.45 (1.25 to 1.64) *** |
| Extremely worried | - | - | - | 2.12 (1.85 to 2.38) *** |

| | | | | |
|---|---|---|---|---|
| Prefer not to say | | - | - | 0.68 (0.27 to 1.09) ** |
| **Region of residence** | - | | | |
| North + Scotland (Reference) | - | - | - | 0.00 |
| Midland + Wales | - | - | - | -0.09 (-0.22 to 0.05) |
| South | - | - | - | -0.04 (-0.19 to 0.11) |
| London | | - | - | -0.08 (-0.23 to 0.08) |
| **Education** | - | | | |
| Degree or equivalent (Reference) | - | - | - | 0.00 |
| Higher education | - | - | - | -0.07 (-0.19 to 0.04) |
| GCE, A level, GCSE or equivalent | - | - | - | -0.07 (-0.36 to 0.21) |
| Other/no qualification | | - | - | -0.42 (-0.76 to -0.09) * |
| **Employment** | - | | | |
| Employed (Reference) | - | - | - | 0.00 |
| Not in labor force | - | - | - | 0.32 (0.17 to 0.46) *** |
| Unemployed | - | - | - | 0.36 (0.12 to 0.59) ** |
| Other/Prefer not to say | | - | - | 0.48 (-0.02 to 0.99) |

* $P < 0.05$; ** $P < 0.01$; *** $P < 0.001$. [a] : adjusted for age, sex, and previous COVID infection; [b] : adjusted for age, sex, previous COVID infection, marital/partnership status, and household generational structure; [c] : adjusted for age, sex, previous COVID infection, marital/partnership status, household generational structure, financial concerns, region of residence, education, and employment status.

Table S4. Complete case analysis: weighted logistic regression analysis across different models using 21 disaggregated ethnic groups (N=13660).

| Variable | Model 1 (without adjusted for any covariate) | Model 2 (adjusted on individual level [a]) | Model 3 (adjusted on individual, and social and household levels [b]) | Model 4 (adjusted on individual, social and household, and socio-economic and environmental levels [c]) |
|---|---|---|---|---|
| | Odds ratio (95% CI) | Odds ratio (95% CI) | Odds ratio (95% CI) | Odds ratio (95% CI) |
| **Ethnicity** | | | | |
| White British (Reference) | 1.00 | 1.00 | 1.00 | 1.00 |
| Asian: Bangladeshi | 1.46 (1.02 to 2.07) * | 1.08 (0.75 to 1.55) | 1.21 (0.84 to 1.75) | 0.87 (0.58 to 1.31) |
| Asian: Chinese | 1.26 (0.93 to 1.69) | 0.94 (0.68 to 1.30) | 1.04 (0.75 to 1.44) | 0.90 (0.63 to 1.28) |
| Asian: Indian | 1.30 (1.07 to 1.58) ** | 1.10 (0.90 to 1.35) | 1.34 (1.08 to 1.65) ** | 1.16 (0.93 to 1.45) |
| Asian: Pakistani | 1.71 (1.34 to 2.18) *** | 1.38 (1.07 to 1.79) * | 1.64 (1.27 to 2.12) *** | 1.29 (0.98 to 1.70) |
| Any other Asian background | 1.41 (1.05 to 1.90) * | 1.14 (0.85 to 1.52) | 1.33 (0.96 to 1.85) | 1.13 (0.78 to 1.62) |
| Black: African | 1.29 (1.02 to 1.63) * | 0.99 (0.77 to 1.29) | 1.11 (0.86 to 1.44) | 0.93 (0.69 to 1.24) |
| Black: Caribbean | 1.75 (1.29 to 2.39) *** | 1.57 (1.13 to 2.20) ** | 1.51 (1.09 to 2.09) * | 1.33 (0.94 to 1.88) |
| Any other Black/African/Caribbean background | 1.57 (0.95 to 2.59) | 1.24 (0.76 to 2.02) | 1.22 (0.70 to 2.12) | 1.06 (0.64 to 1.77) |
| Mixed: White and Asian | 1.40 (1.04 to 1.89) * | 1.02 (0.75 to 1.39) | 1.07 (0.79 to 1.45) | 0.92 (0.67 to 1.27) |
| Mixed: White and Black African | 1.70 (1.00 to 2.91) | 1.29 (0.74 to 2.27) | 1.21 (0.72 to 2.04) | 1.10 (0.65 to 1.85) |
| Mixed: White and Black Caribbean | 2.61 (1.85 to 3.68) *** | 1.87 (1.32 to 2.65) *** | 1.79 (1.25 to 2.57) ** | 1.72 (1.16 to 2.54) ** |
| Any other mixed/multiple background | 1.90 (1.38 to 2.63) *** | 1.44 (1.02 to 2.03) * | 1.46 (0.99 to 2.14) | 1.20 (0.80 to 1.81) |
| White: Eastern European | 1.94 (1.37 to 2.75) *** | 1.53 (1.04 to 2.26) * | 1.71 (1.14 to 2.55) ** | 1.61 (1.05 to 2.49) * |
| White: Gypsy/Traveller | 0.79 (0.42 to 1.46) | 0.64 (0.35 to 1.16) | 0.71 (0.41 to 1.23) | 0.61 (0.35 to 1.04) |
| White: Irish | 1.02 (0.63 to 1.62) | 1.02 (0.63 to 1.67) | 1.05 (0.65 to 1.69) | 0.96 (0.60 to 1.52) |
| White: Roma | 0.65 (0.33 to 1.27) | 0.47 (0.23 to 0.95) * | 0.42 (0.21 to 0.84) * | 0.37 (0.18 to 0.75) ** |
| Any other White background | 1.29 (1.04 to 1.60) * | 1.03 (0.82 to 1.29) | 1.04 (0.82 to 1.31) | 0.96 (0.75 to 1.24) |
| Other: Arab | 1.63 (0.96 to 2.77) | 1.25 (0.70 to 2.23) | 1.44 (0.78 to 2.64) | 1.11 (0.63 to 1.96) |
| Any other ethnic group | 2.15 (1.44 to 3.20) *** | 1.85 (1.20 to 2.87) ** | 1.98 (1.29 to 3.05) ** | 1.49 (0.94 to 2.34) |
| Jewish | 0.92 (0.73 to 1.15) | 0.93 (0.73 to 1.19) | 1.07 (0.83 to 1.37) | 0.96 (0.72 to 1.29) |

| | | | | |
|---|---|---|---|---|
| **Age (years old)** | - | | | |
| 18-24 (Reference) | - | 1.00 | 1.00 | 1.00 |
| 25-34 | - | 0.70 (0.53 to 0.94) * | 0.83 (0.61 to 1.12) | 0.76 (0.56 to 1.03) |
| 35-44 | - | 0.61 (0.46 to 0.81) ** | 0.80 (0.59 to 1.08) | 0.74 (0.54 to 1.01) |
| 45-54 | - | 0.50 (0.37 to 0.66) *** | 0.63 (0.47 to 0.86) ** | 0.56 (0.41 to 0.77) *** |
| 55-64 | - | 0.42 (0.31 to 0.56) *** | 0.56 (0.40 to 0.79) ** | 0.52 (0.37 to 0.73) *** |
| 65-75 | | 0.26 (0.19 to 0.35) *** | 0.31 (0.22 to 0.45) *** | 0.32 (0.22 to 0.47) *** |
| **Sex** | - | | | |
| Male (Reference) | - | 1.00 | 1.00 | 1.00 |
| Female | | 1.33 (1.17 to 1.52) *** | 1.26 (1.10 to 1.45) ** | 1.14 (0.99 to 1.32) |
| **Previous COVID infection** | - | | | |
| Ever had positive results (Reference) | - | 1.00 | 1.00 | 1.00 |
| Never tested | - | 0.94 (0.72 to 1.23) | 0.89 (0.67 to 1.17) | 0.88 (0.66 to 1.17) |
| Never had positive results | - | 0.97 (0.75 to 1.26) | 0.92 (0.70 to 1.20) | 0.89 (0.68 to 1.17) |
| Other/Prefer not to say | | 1.41 (0.84 to 2.37) | 1.30 (0.79 to 2.14) | 1.07 (0.63 to 1.81) |
| **Marital/Partnership status** | - | | | |
| Married/Civil (Reference) | - | - | 1.00 | 1.00 |
| Not married/not civil | - | - | 2.03 (1.71 to 2.42) *** | 1.86 (1.55 to 2.22) *** |
| Separated/Divorced/Dissolved | - | - | 2.59 (2.07 to 3.23) *** | 2.38 (1.87 to 3.03) *** |
| Widowed/Partner died | - | - | 3.15 (2.20 to 4.53) *** | 3.04 (2.07 to 4.46) *** |
| Prefer not to say | | - | 1.82 (1.05 to 3.14) * | 1.55 (0.87 to 2.74) |
| **Number of generations living in household** | - | | | |
| One generation (Reference) | - | - | 1.00 | 1.00 |
| Two generations | - | - | 0.98 (0.84 to 1.15) | 0.91 (0.77 to 1.07) |
| Three generations | - | - | 1.14 (0.78 to 1.68) | 1.01 (0.69 to 1.48) |
| Other/Prefer not to say | | - | 1.38 (0.73 to 2.61) | 1.26 (0.65 to 2.45) |
| **Worrying about financial situation** | - | | | |
| Not at all worried (Reference) | - | - | - | 1.00 |
| Somewhat worried | - | - | - | 2.66 (2.24 to 3.16) *** |
| Very worried | - | - | - | 4.67 (3.55 to 6.15) *** |
| Extremely worried | - | - | - | 7.29 (5.41 to 9.83) *** |

| | | | | |
|---|---|---|---|---|
| Prefer not to say | | - | - | 2.41 (1.47 to 3.95) *** |
| **Region of residence** | - | | | |
| North + Scotland (Reference) | - | - | - | 1.00 |
| Midland + Wales | - | - | - | 0.91 (0.76 to 1.09) |
| South | - | - | - | 0.97 (0.79 to 1.19) |
| London | | - | - | 1.00 (0.81 to 1.22) |
| **Education** | - | | | |
| Degree or equivalent (Reference) | - | - | - | 1.00 |
| Higher education | - | - | - | 0.93 (0.80 to 1.08) |
| GCE, A level, GCSE or equivalent | - | - | - | 0.84 (0.59 to 1.20) |
| Other/no qualification | | - | - | 0.64 (0.40 to 1.04) |
| **Employment** | - | | | |
| Employed (Reference) | - | - | - | 1.00 |
| Not in labor force | - | - | - | 1.38 (1.13 to 1.68) ** |
| Unemployed | - | - | - | 1.39 (1.07 to 1.81) * |
| Other/Prefer not to say | | - | - | 1.40 (0.79 to 2.46) |

* $P < 0.05$; ** $P < 0.01$; *** $P < 0.001$. [a] : adjusted for age, sex, and previous COVID infection; [b] : adjusted for age, sex, previous COVID infection, marital/partnership status, and household generational structure; [c] : adjusted for age, sex, previous COVID infection, marital/partnership status, household generational structure, financial concerns, region of residence, education, and employment status.

Table S5: Weight adjusted differences in loneliness across different models using 6 aggregated ethnic groups (N=13967).

| Variable | Model 1 (without adjusted for any covariate) | Model 2 (adjusted on individual level [a]) | Model 3 (adjusted on individual, and social and household levels [b]) | Model 4 (adjusted on individual, social and household, and socio-economic and environmental levels [c]) |
|---|---|---|---|---|
| | Odds ratio (95% CI) | Odds ratio (95% CI) | Odds ratio (95% CI) | Odds ratio (95% CI) |
| **Ethnicity** | | | | |
| White British (Reference) | 1.00 | 1.00 | 1.00 | 1.00 |
| White minority | 1.39 (1.16 to 1.65) *** | 1.13 (0.94 to 1.37) | 1.18 (0.97 to 1.44) | 1.10 (0.89 to 1.36) |
| Mixed or multiple ethnic groups | 1.92 (1.59 to 2.32) *** | 1.41 (1.15 to 1.73) ** | 1.41 (1.14 to 1.74) ** | 1.27 (1.01 to 1.58) * |
| Asian | 1.47 (1.28 to 1.69) *** | 1.19 (1.03 to 1.38) * | 1.39 (1.19 to 1.62) *** | 1.16 (0.98 to 1.37) |
| Black | 1.46 (1.21 to 1.75) *** | 1.18 (0.96 to 1.44) | 1.25 (1.02 to 1.52) * | 1.07 (0.85 to 1.34) |
| Other ethnic groups | 1.68 (1.23 to 2.28) ** | 1.51 (1.08 to 2.11) * | 1.63 (1.17 to 2.27) ** | 1.29 (0.91 to 1.83) |
| **Age (years old)** | - | | | |
| 18-24 (Reference) | - | 1.00 | 1.00 | 1.00 |
| 25-34 | - | 0.71 (0.54 to 0.94) * | 0.84 (0.63 to 1.12) | 0.77 (0.57 to 1.04) |
| 35-44 | - | 0.62 (0.47 to 0.82) ** | 0.81 (0.60 to 1.09) | 0.75 (0.55 to 1.02) |
| 45-54 | - | 0.50 (0.38 to 0.66) *** | 0.64 (0.47 to 0.87) ** | 0.57 (0.42 to 0.78) *** |
| 55-64 | - | 0.43 (0.32 to 0.57) *** | 0.57 (0.41 to 0.79) ** | 0.53 (0.38 to 0.74) *** |
| 65-75 | | 0.26 (0.19 to 0.35) *** | 0.31 (0.22 to 0.45) *** | 0.33 (0.23 to 0.47) *** |
| **Sex** | - | | | |
| Male (Reference) | - | 1.00 | 1.00 | 1.00 |
| Female | | 1.33 (1.17 to 1.52) *** | 1.26 (1.10 to 1.45) ** | 1.14 (0.99 to 1.32) |
| **Previous COVID infection** | - | | | |
| Ever had positive results (Reference) | - | 1.00 | 1.00 | 1.00 |
| Never tested | - | 0.95 (0.73 to 1.24) | 0.90 (0.68 to 1.17) | 0.88 (0.67 to 1.17) |
| Never had positive results | - | 0.97 (0.75 to 1.25) | 0.91 (0.70 to 1.19) | 0.89 (0.68 to 1.16) |
| Other/Prefer not to say | | 1.50 (0.90 to 2.49) | 1.38 (0.85 to 2.24) | 1.14 (0.68 to 1.91) |
| **Marital/Partnership status** | - | | | |

| | | | | |
|---|---|---|---|---|
| Married/Civil (Reference) | - | - | 1.00 | 1.00 |
| Not married/not civil | - | - | 2.00 (1.68 to 2.37) *** | 1.83 (1.53 to 2.18) *** |
| Separated/Divorced/Dissolved | - | - | 2.54 (2.04 to 3.16) *** | 2.33 (1.85 to 2.96) *** |
| Widowed/Partner died | - | - | 3.24 (2.27 to 4.63) *** | 3.10 (2.13 to 4.52) *** |
| Prefer not to say | | - | 1.83 (1.08 to 3.08) * | 1.57 (0.91 to 2.70) |
| **Number of generations living in household** | - | | | |
| One generation (Reference) | - | - | 1.00 | 1.00 |
| Two generations | - | - | 0.99 (0.85 to 1.16) | 0.92 (0.78 to 1.08) |
| Three generations | - | - | 1.14 (0.79 to 1.66) | 1.00 (0.69 to 1.45) |
| Other/Prefer not to say | | - | 1.35 (0.72 to 2.56) | 1.20 (0.62 to 2.33) |
| **Worrying about financial situation** | - | | | |
| Not at all worried (Reference) | - | - | - | 1.00 |
| Somewhat worried | - | - | - | 2.65 (2.24 to 3.15) *** |
| Very worried | - | - | - | 4.75 (3.63 to 6.22) *** |
| Extremely worried | - | - | - | 7.45 (5.55 to 9.99) *** |
| Prefer not to say | | - | - | 2.61 (1.64 to 4.16) *** |
| **Region of residence** | - | | | |
| North + Scotland (Reference) | - | - | - | 1.00 |
| Midland + Wales | - | - | - | 0.91 (0.76 to 1.09) |
| South | - | - | - | 0.96 (0.78 to 1.17) |
| London | | - | - | 0.97 (0.79 to 1.18) |
| **Education** | - | | | |
| Degree or equivalent (Reference) | - | - | - | 1.00 |
| Higher education | - | - | - | 0.94 (0.81 to 1.00) |
| GCE, A level, GCSE or equivalent | - | - | - | 0.80 (0.56 to 1.13) |
| Other/no qualification | | - | - | 0.69 (0.44 to 1.09) |
| **Employment** | - | | | |
| Employed (Reference) | - | - | - | 1.00 |
| Not in labor force | - | - | - | 1.38 (1.13 to 1.67) ** |
| Unemployed | - | - | - | 1.34 (1.03 to 1.75) * |
| Other/Prefer not to say | | - | - | 1.40 (0.82 to 2.41) |

* P < 0.05; ** P < 0.01; *** P < 0.001. [a] : adjusted for age, sex, and previous COVID infection; [b] : adjusted for age, sex, previous COVID infection, marital/partnership status, and household generational structure; [c] : adjusted for age, sex, previous COVID infection, marital/partnership status, household generational structure, financial concerns, region of residence, education, and employment status.

Table S6. Variance Inflation Factor (VIF) scores of independent variables.

| Variable (Continuous) | VIF |
|---|---|
| **Ethnicity** | |
| White British | Reference |
| Asian: Bangladeshi | 1.15 |
| Asian: Chinese | 1.20 |
| Asian: Indian | 1.34 |
| Asian: Pakistani | 1.25 |
| Any other Asian background | 1.21 |
| Black: African | 1.32 |
| Black: Caribbean | 1.17 |
| Any other Black/African/Caribbean background | 1.05 |
| Mixed: White and Asian | 1.15 |
| Mixed: White and Black African | 1.05 |
| Mixed: White and Black Caribbean | 1.10 |
| Any other mixed/multiple background | 1.11 |
| White: Eastern European | 1.11 |
| White: Gypsy/Traveller | 1.39 |
| White: Irish | 1.03 |
| White: Roma | 1.11 |
| Any other White background | 1.15 |
| Other: Arab | 1.06 |
| Any other ethnic group | 1.08 |
| Jewish | 1.19 |
| **Age (years old)** | |
| 18-24 | Reference |
| 25-34 | 2.28 |
| 35-44 | 2.36 |
| 45-54 | 2.21 |
| 55-64 | 2.07 |
| 65-75 | 2.66 |
| **Sex** | |
| Male | Reference |
| Female | 1.05 |
| **Infection** | |
| Ever had positive results | Reference |
| Never tested | 3.07 |
| Never had positive results | 3.00 |
| Other/Prefer not to say | 1.21 |
| **Marital/Partnership status** | |
| Married/Civil | Reference |
| Not married/not civil | 1.63 |

| | |
|---|---|
| Separated/Divorced/Dissolved | 1.12 |
| Widowed/Partner died | 1.10 |
| Prefer not to say | 1.07 |
| **Number of generations living in household** | |
| One generation | Reference |
| Two generations | 1.32 |
| Three generations | 1.11 |
| Other/Prefer not to say | 1.05 |
| **Worrying about financial situation** | |
| Not at all worried | Reference |
| Somewhat worried | 1.47 |
| Very worried | 1.34 |
| Extremely worried | 1.26 |
| Prefer not to say | 1.09 |
| **Region** | |
| North + Scotland | Reference |
| Midland + Wales | 1.53 |
| South | 1.37 |
| London | 1.62 |
| Education | |
| Degree or equivalent | Reference |
| Higher education | 1.17 |
| GCE, A level, GCSE or equivalent | 1.45 |
| Other/no qualification | 1.04 |
| Employment | |
| Employed | Reference |
| Not in labor force | 1.44 |
| Unemployed | 1.08 |
| Other/Prefer not to say | 1.06 |
| Mean VIF | 1.40 |
